# Country-level Association of Socioeconomic, Environmental and Healthcare-Related Factors with the Disease-Burden and Mortality Rate of COVID-19

**DOI:** 10.1101/2021.08.22.21262449

**Authors:** Tasnuva Chowdhury, Upal Mahbub, Tauhidur Rahman

**Affiliations:** University of California San Diego, San Diego, CA, 92093.; Qualcomm Technologies Inc., San Diego, CA, 92121.; College of Information and Computer Sciences, University of Massachusetts Amherst, MA 01003.

**Keywords:** COVID-19, SARS-CoV-2, disease burden, mortality rate, healthcare infrastructure, country-level analysis, multiple linear regression, backward elimination

## Abstract

**Background:** COVID-19 pandemic is rapidly expanding throughout the world right now. Caused by a novel strain of the coronavirus, the manifestation of this pandemic shows a unique level of disease burden and mortality rate in different countries.

**Objective:** In this paper, we investigated the effects of several socioeconomic, environmental, and healthcare-related factors on the disease burden and mortality rate of COIVID-19 across countries. Our main objective is to provide a macro-level understanding of the most influential socioeconomic, environmental, and healthcare-related factors associated with the disease burden and mortality rate metrics without human bias.

**Methods:** We developed a multiple linear regression model using backward elimination to find the best fitting between reported death and cases across countries for country-level aggregated independent factors keeping COVID-19 test statistic in consideration. Notably, the method requires minimum human intervention and handles confounding effects intrinsically.

**Results:** Our results show that while the COVID-19 pandemic is seemingly spreading more rapidly in economically affluent countries, it Is more deadly in countries with inadequate healthcare infrastructure, lower capacity of handling epidemics, and lower allocation of the healthcare budget. We also did not find evidence of any association between environmental factors and COVID-19.

**Conclusion:** We took the number of tests performed into account and normalized the case and mortality counts based on the cumulative distribution of cases across days. Our analysis of the standardized factors provides both the direction and relative importance of different factors leading to several compelling insights into the most influential socioeconomic and healthcare infrastructure-related factors from a country-level view.

## Introduction

As 2019 was coming to an end, a deadly virus emerged in Wuhan, Hubei Province of China. Patients got admitted to the hospitals with pneumonia-like symptoms, and by 03 January 2020, 11 patients became critically ill of the 44 reported cases in Wuhan City [1]. The virus, SARS-CoV-2, got its name because of its genetic resemblance with beta coronavirus lineage that caused the 2003 SARS outbreak, and the disease is named COVID-19 [2]. Soon, the infection spread all over the world mostly through human to human transmission, and on 11 March 2020, the World Health Organization (WHO) declared COVID-19 as a worldwide pandemic based on its severity and spread (118,000 cases and 4,291 deaths worldwide) [3] [4]. As of 01 May 2020, when we performed this experiment, COVID-19 caused 3,394,153 cases and 239,447 deaths worldwide [5].

SARS-CoV-2 can spread by silent carriers to the community, and it has a significantly higher respiratory load than SARS-CoV-1 [6] [7]. It can spread directly or indirectly through respiratory droplets released in the air or landed on high-touch surfaces [8]. Apart from encouraging people to maintain hand hygiene and social distancing, governments of the most affected countries are strictly prohibiting large gatherings, enforcing lockdown, canceling most international flights, advising the local restaurants and small businesses to stay closed or operate at a limited business hour, and increasing their capability to test the population for COVID-19 [9] [10] [11].

There has been a significant research effort from public health and many other communities to investigate the factors associated with COVID-19 [12] [13] [14] [15]. For example, in [12], the authors tried to characterize the COVID-19 spread patterns among different age groups considering four different social contact settings. In [14], the authors did a cohort study over 72000 individuals of 35 to 41 years of age to investigate the association of BCG vaccination with COVID-19 severity, which came out to be inconclusive. In an earlier study [15], the authors claimed a significant association of the disease with the temperature. Several studies found an association of COVID-19 severity with obesity, age, smoking habit, and chronic medical conditions such as diabetes and hypertension [16] [17] [18] [19] [20]. Other studies found relations with low levels of surfactants in lungs, genetic predisposition, the occurrence of cytokine storm, and the amount of the virus load [21] [22].

However, these works are mostly patient-level analysis not considering health-infrastructure related factors – which, according to our study, seem to have a confounding impact on country-level aggregates of many of these factors. In [23], the authors did a country-level aggregated research on COVID-19 to find the weather, socioeconomic and geographic factors associated with the cases and deaths of the disease. The authors claimed that low temperature, low humidity, and higher altitude play a role in the rapid spread of the pandemic. However, recent studies found inconclusive relation between temperature and COVID-19 in different parts of the world [24] [25].

In this paper, we concentrated on a country-level aggregate to get a macro-level understanding of why the disease is affecting different countries in different ways. To make a fair comparison, we performed a unique normalization considering a 30-day time window at dynamic start dates for various countries. This normalization ensured that the dependent variables represent the rate of changes of the disease burden and mortality within the same time window for every country. The major highlights of this work are

1. We determined the best-fitted models for disease burden and mortality due to COVID-19 utilizing cross-correlation coefficients for initial factor selection and backward elimination methods. We applied the technique on a wide range of factors covering a broad scope. That way, the confounding effects are intrinsically handled, and human choices do not bias the best-fitted models.
2. We demonstrated that the countries with more burden of the disease are not always the ones with the highest mortality rate.
3. Our analysis shows that a country’s healthcare infrastructure plays a determining role when it comes to the effect of COVID-19. Also, environmental factors are absent in the best-fitted models for disease burden and mortality rate.
4. Our results show insightful relation of the disease-burden and mortality rate metrics with GDP, healthcare infrastructure, net-migration, phones per 1000 population, and few other country-level factors considering the number of tests performed.

In the next section, we discuss the research methodology with details on the datasets, pre-processing steps, and initial factor selection. The results and analysis section follows after that. The final section contains discussions on the findings, limitations of this work, and future extension ideas.

## Methods

### Dataset and pre-processing

The full list of variables, their definition, and data sources are provided in

**Table I**. We performed the following pre-processing steps before the analysis:

**Table I:**
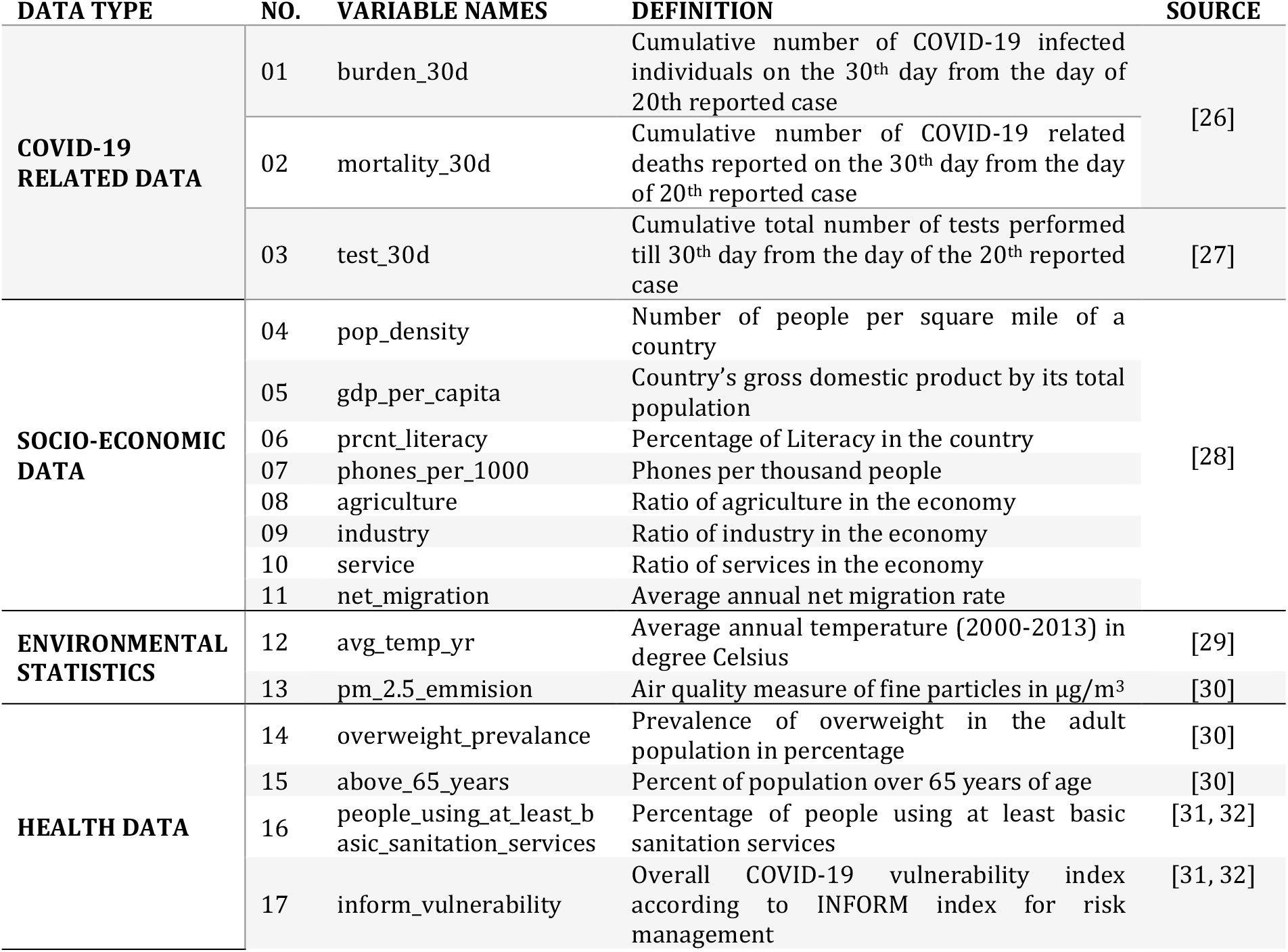
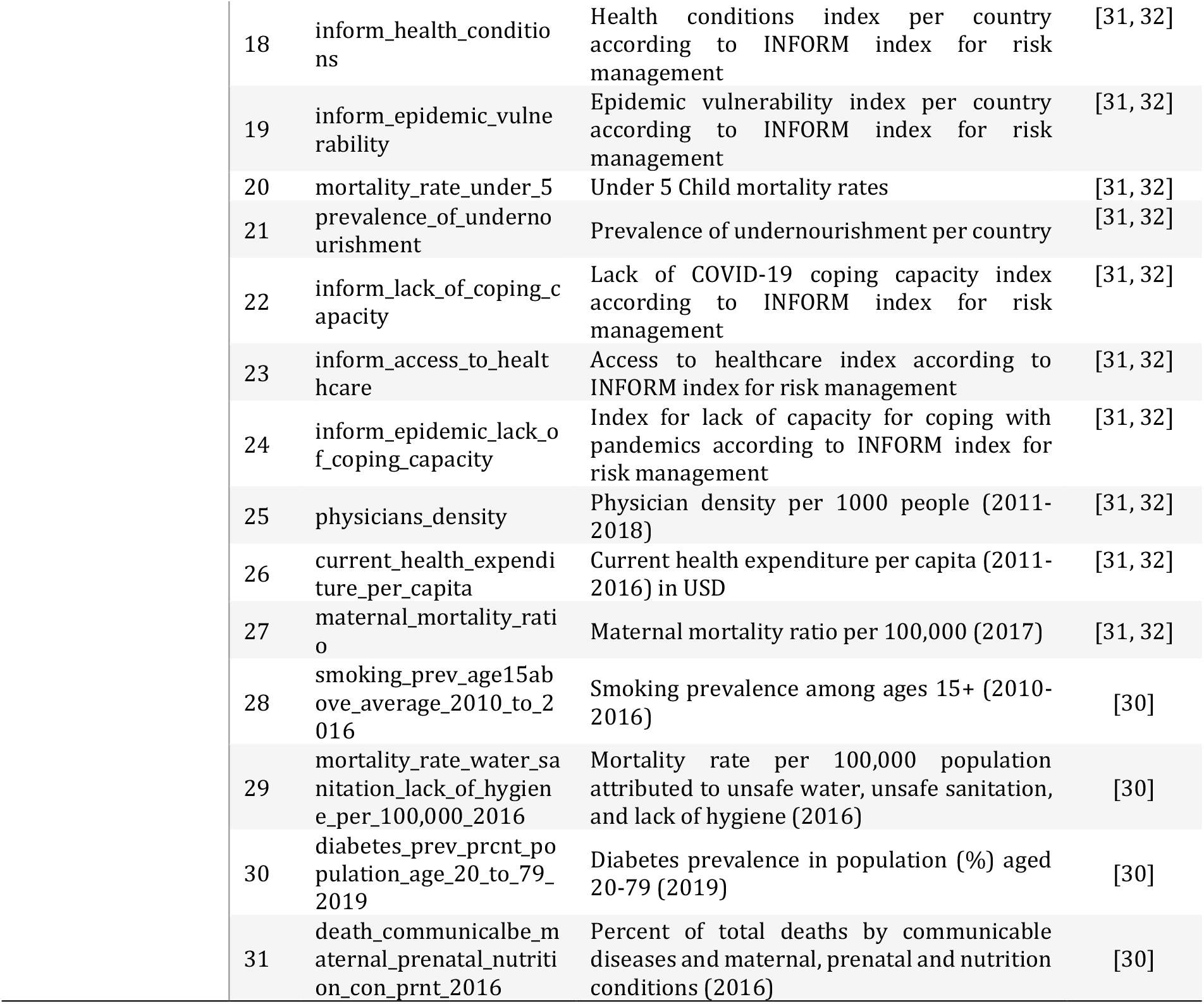
List of variables used in the analysis along with variable definitions and sources.

- The counts of cases and deaths across the countries were standardized. We identified the day when a country reached its first 20 cases as the Day 1 for that country. Starting from Day 1, we calculated the Day 30 for each country and considered the numbers of cases and deaths for Day 30 as the standardized data of any country. Considering the variation in Day 1 for different countries, we found 112 countries with cases and deaths information for Day 30 at the time of the analysis.
- We removed all the countries with any missing values in COVID-19 cases, deaths, and tests. The final curated dataset has a total of 60 countries.

We considered the variables *burden_30d* and *mortality_30d*, representing the disease-burden and mortality for the disease in the 30 days window, as the dependent variables. The *test_30d* is considered by default for all models to nullify the impact of the different number of tests performed across various countries. When regressing the model for *mortality_30d*, the variable *burden_30d* is considered as a confounding factor to ensure that the impact of the disease-burden is considered when modeling the mortality rate.

We consider a health system to comprise of resources, institutions, and organizations delivering services to improve the health of a target population, including public, private, and informal sectors [33] [34]. Since health systems can vary substantially from country to country, we picked the indicators described under Health Data in **Table I**. A few of these factors related to lack of coping capacity and vulnerability are obtained from the INFORM index for risk management [31]. Other factors such as physician density, health expenditure, and mortality rates are commonly used as indicators for country-level health system performance [35] [30].

### Finding Best-Fitted Model

We took a two-step approach to find the best-fitted model. First, we performed Pearson correlation tests to find the pair-wise correlation coefficients for all the other factors with the *burden_30d* and *mortality_30d* data. Then, for initial factor selection, we filtered out any factor with p-values greater than 0.1 and a magnitude less than 0.2 from further analysis. Next, we performed multiple linear regression iteratively with backward elimination to find the best-fitted model for disease burden and mortality. Backward elimination is a well-known variable selection technique, as discussed in [36]. Since our primary goal is to find the statistically significant parameters to fit the dependent variables best, we resorted to multiple linear regression analysis as a fast and baseline model. It is indeed possible to extend the experiment into building more sophisticated regression models such as Poisson or Negative Binomial models. However, as explained in chapter 3 of [37], the most statistically significant parameters of the multiple linear regression are the same as the most statistically significant parameters of the Poisson and the Negative Binomial Regression Models.

## Results and Discussion

### Initial Factor Selection

We show the absolute values of the correlation coefficients in **Figure 1**, where the yellow bars represent coefficients values above the 0.2 thresholds.

**Figure 1:**
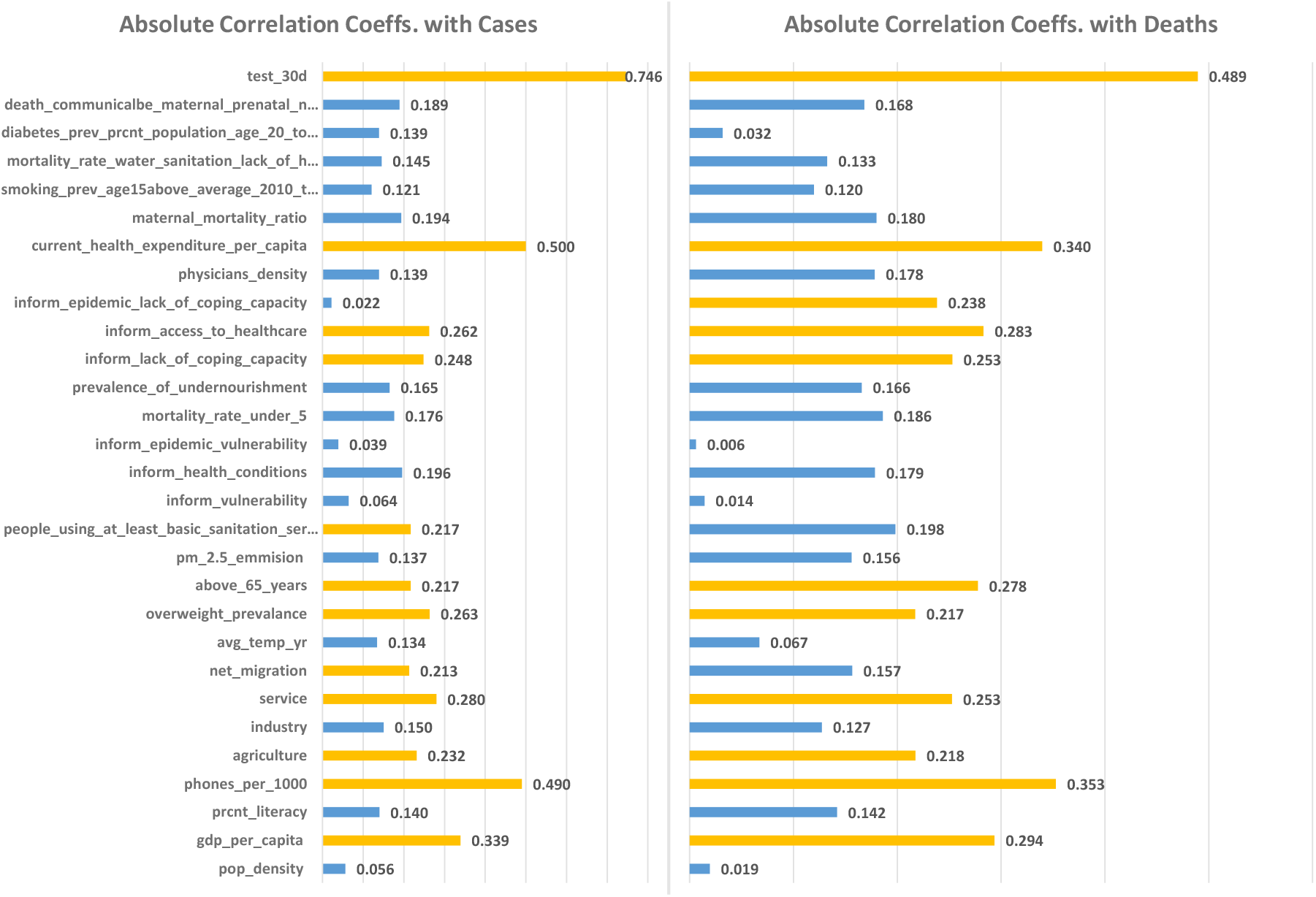
Absolute values of Correlation Coefficients between all factors and burden_30d (left) and mortality_30d (right).

After the correlation-based selection, we ended up with 12 factors correlated with *burden_30d* and 11 factors with *mortality_30d*. As mentioned earlier, for *mortality_30d*, the *burden_30d* variable is considered as an additional factor to nullify the confounding effect of the disease burden when performing multivariate analysis. We present the descriptive statistics for *burden_30d, mortality_30d*, and all the initially selected factors in Table II.

**Table II:** Descriptive Statistics of the initially Selected Factors.

| VARIABLE NAMES | MEAN | STD | MIN | MAX | IQR | MEDIAN | KURTOSIS | SKEWNESS |
| --- | --- | --- | --- | --- | --- | --- | --- | --- |
| **burden_30d** | 11709.1 | 25936.54 | 47 | 161679 | 6494.8 | 3257.5 | 19.70 | 4.10 |
| **mortality_30d** | 491.42 | 1265.77 | 0 | 6803 | 239 | 69 | 14.63 | 3.77 |
| **gdp_per_capita** | 15183.33 | 12102.75 | 1000.0 | 55100.0 | 21450.0 | 10900.0 | 0.38 | 0.90 |
| **phones_per_1000** | 306.66 | 208.06 | 2.70 | 898.00 | 335.80 | 288.60 | -0.38 | 0.41 |
| **agriculture** | 0.08 | 0.08 | 0.01 | 0.40 | 0.08 | 0.05 | 4.58 | 1.85 |
| **service** | 0.63 | 0.10 | 0.37 | 0.86 | 0.12 | 0.64 | 0.25 | -0.47 |
| **net_migration** | 0.45 | 2.51 | -8.58 | 8.97 | 2.13 | 0.00 | 3.93 | -0.04 |
| **overweight_prevalance** | 51.92 | 13.69 | 16.78 | 66.62 | 5.67 | 56.83 | 0.92 | -1.53 |
| **above_65_years** | 12.52 | 6.00 | 2.11 | 25.54 | 11.05 | 13.97 | -1.21 | -0.06 |
| **people_using_at_least_basic_sanitation_services** | 89.81 | 15.35 | 29.05 | 100.00 | 11.03 | 97.66 | 4.22 | -2.08 |
| inform_lack_of_coping_capacity | 3.14 | 1.44 | 0.90 | 6.50 | 2.43 | 2.90 | -0.86 | 0.43 |
| inform_access_to_healthcare | 2.51 | 1.96 | 0.20 | 7.70 | 3.03 | 2.00 | -0.54 | 0.71 |
| inform_epidemic_lack_of_coping_capacity | 3.24 | 1.68 | 1.10 | 8.20 | 2.00 | 2.90 | 0.94 | 0.98 |
| current_health_expenditure_per_capita | 2495.96 | 2123.66 | 90.60 | 9869.74 | 3276.96 | 1912.60 | 1.39 | 1.19 |
| test_30d | 121671.02 | 215769.65 | 2517.0 | 1004719.0 | 79402.50 | 48126.5 | 9.43 | 3.11 |

It is important to note that the mean for each variable is varying in a wide range because of the different units. Hence, we normalized each variable with their corresponding mean and standard deviation and provided the standardized beta coefficients for each variable after regression.

### Model Fitting

The result tables for initial multiple linear regression for *cases* and *deaths* are presented in **Table III** and **Table IV**. We performed backward elimination for factor selection and dropped the factor with the highest p-value before performing the next multivariate regression. For example, the variable above_65_years (p-value = 0.93) is to be dropped based on the results presented in **Table III**.

**Table III:** Results of multivariate regression between burden_30d and all the 12 selected variables.

| Regression Statistics |  |
| --- | --- |
| Multiple R | 0.8365 |
| R Square | 0.6998 |
| Adjusted R Square | 0.6231 |
| Standard Error | 15922.7567 |
| Observations | 60.0000 |

|  | Standardized Coefficients | Standard Error | t Stat | P-value | Lower 95% | Upper 95% |
| --- | --- | --- | --- | --- | --- | --- |
| Intercept | 7.67E-17 | 0.08 | 0.00 | 1.00 | -0.16 | 0.16 |
| gdp_per_capita | -0.21 | 0.28 | -0.73 | 0.47 | -0.78 | 0.36 |
| phones_per_1000 | 0.21 | 0.24 | 0.89 | 0.38 | -0.27 | 0.69 |
| agriculture | 0.21 | 0.15 | 1.42 | 0.16 | -0.09 | 0.50 |
| service | 0.14 | 0.14 | 0.97 | 0.34 | -0.15 | 0.42 |
| net_migration | -0.20 | 0.13 | -1.49 | 0.14 | -0.46 | 0.07 |
| overweight_prevalance | 0.11 | 0.13 | 0.85 | 0.40 | -0.16 | 0.39 |
| above_65_years | 0.02 | 0.18 | 0.09 | 0.93 | -0.34 | 0.37 |
| people_using_at_least_basic_sanitation_services | 0.11 | 0.21 | 0.55 | 0.59 | -0.31 | 0.54 |
| inform_lack_of_coping_capacity | -0.03 | 0.22 | -0.13 | 0.90 | -0.46 | 0.41 |
| inform_access_to_healthcare | 0.44 | 0.24 | 1.84 | 0.07 | -0.04 | 0.93 |
| current_health_expenditure_per_capita | 0.63 | 0.26 | 2.45 | 0.02 | 0.11 | 1.16 |
| test_30d | 0.60 | 0.10 | 5.98 | 0.00 | 0.40 | 0.81 |

**Table IV:** Results of multivariate regression between mortality_30d and all the 12 selected variables.

| Regression Statistics |  |
| --- | --- |
| Multiple R | 0.8960 |
| R Square | 0.8028 |
| Adjusted R Square | 0.7524 |
| Standard Error | 629.8497 |
| Observations | 60.0000 |

|  | Standardized Coefficients | Standard Error | t Stat | P-value | Lower 95% | Upper 95% |
| --- | --- | --- | --- | --- | --- | --- |
| Intercept | 2.97E-17 | 0.06 | 0.00 | 1.00 | -0.13 | 0.13 |
| gdp_per_capita | 0.532 | 0.22 | 2.45 | 0.02 | 0.09 | 0.97 |
| phones_per_1000 | -0.265 | 0.20 | -1.35 | 0.18 | -0.66 | 0.13 |
| agriculture | -0.046 | 0.12 | -0.39 | 0.69 | -0.28 | 0.19 |
| service | -0.002 | 0.10 | -0.02 | 0.98 | -0.20 | 0.20 |
| overweight_prevalance | -0.102 | 0.10 | -1.04 | 0.31 | -0.30 | 0.10 |
| above_65_years | 0.104 | 0.15 | 0.70 | 0.49 | -0.20 | 0.40 |
| inform_lack_of_coping_capacity | 0.018 | 0.16 | 0.11 | 0.91 | -0.30 | 0.34 |
| inform_access_to_healthcare | -0.373 | 0.18 | -2.12 | 0.04 | -0.73 | -0.02 |
| inform_epidemic_lack_of_copin<br>g_capacity | 0.138 | 0.07 | 1.90 | 0.06 | -0.01 | 0.28 |
| current_health_expenditure_pe<br>r_capita | -0.732 | 0.22 | -3.33 | 0.00 | -1.17 | -0.29 |
| burden_30d | 1.210 | 0.12 | 10.40 | 0.00 | 0.98 | 1.44 |
| test_30d | -0.263 | 0.11 | -2.50 | 0.02 | -0.47 | -0.05 |

We compared the adjusted R-square values for consecutive models to see if the latest model fits the dependent variable better (higher adjusted R-square denotes better fit).

In **Figure 2**, the changes in adjusted R-square values at different iteration steps are presented for both cases (left) and depth (right) estimation. The variable with the maximum p-value at any state is shown along the x-axis for that state. It can be seen that the adjusted R-square value reaches a peak after a few iterations before going down again. The models associated with the adjusted R-square peaks are the best-fitted models obtained from the regression analysis (**Table V** and **Table VI**).

**Figure 2:**
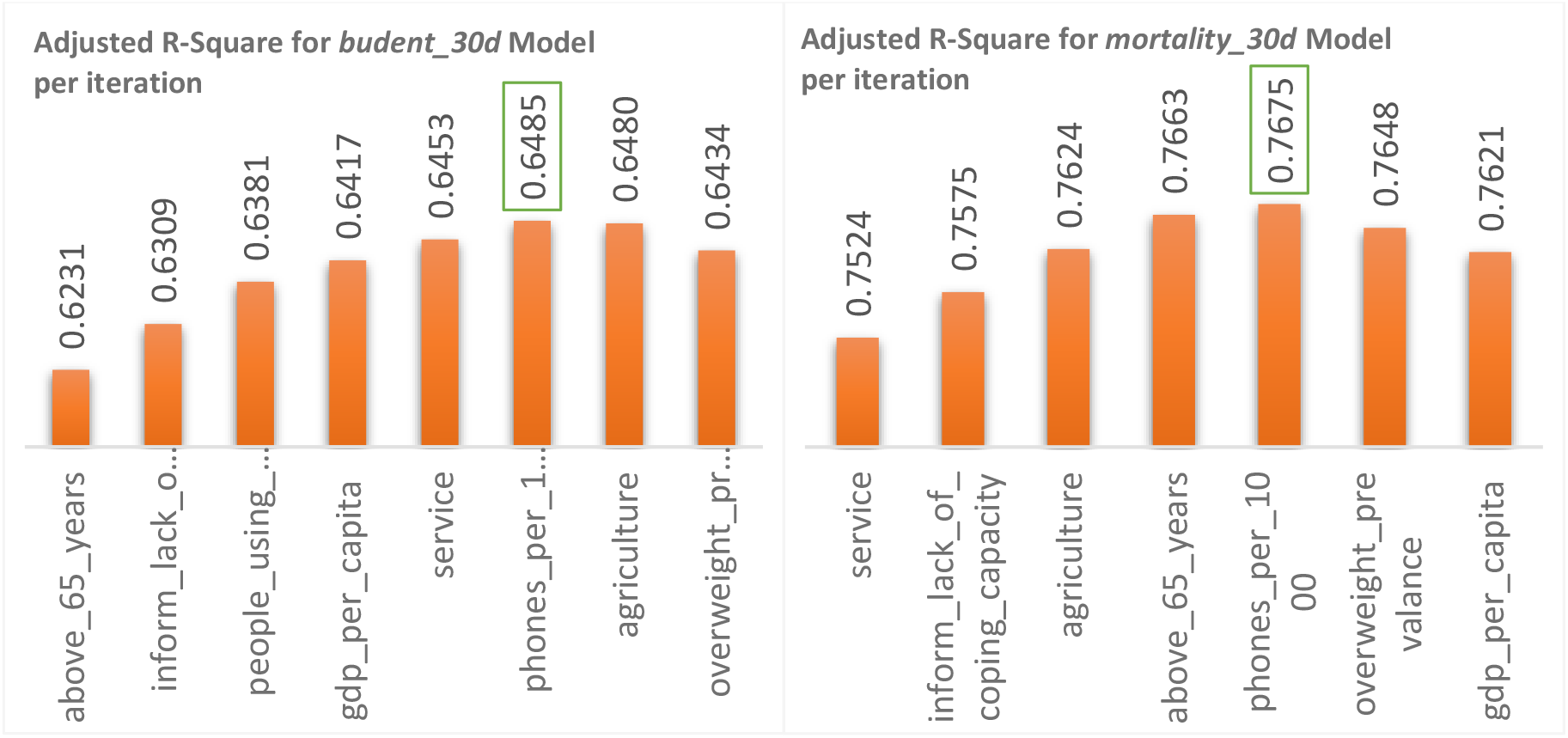
Changes in adjusted R-Square values when performing backward elimination for burden_30d (left) and mortality_30d (right) models with the selected factors. The variables along the x-axis are the ones that have the highest p-value at that instance and were dropped before doing the next multivariate regression.

**Table V:** Best fitted model statistics for cases.

| *Regression Statistics* |  |
| --- | --- |
| Multiple R | 0.8308 |
| R Square | 0.6902 |
| Adjusted R Square | 0.6485 |
| Standard Error | 0.5929 |
| Observations | 60.0000 |

|  | *Standardized Coefficients* | *Standard Error* | *t Stat* | *P-value* | *Lower 95%* | *Upper 95%* |
| --- | --- | --- | --- | --- | --- | --- |
| Intercept | 8.26E-19 | 0.08 | 1.08E-17 | 1 | -0.15 | 0.15 |
| phones_per_1000 | 0.23 | 0.22 | 1.03 | 0.31 | -0.22 | 0.67 |
| agriculture | 0.18 | 0.13 | 1.41 | 0.16 | -0.07 | 0.43 |
| net_migration | -0.25 | 0.11 | -2.34 | 0.02 | -0.46 | -0.04 |
| overweight_prevalance | 0.18 | 0.10 | 1.72 | 0.09 | -0.03 | 0.38 |
| inform_access_to_health care | 0.37 | 0.16 | 2.28 | 0.03 | 0.04 | 0.70 |
| current_health_expenditure_per_capita | 0.55 | 0.19 | 2.91 | 0.01 | 0.17 | 0.93 |
| test_30d | 0.60 | 0.09 | 6.79 | 0.00 | 0.42 | 0.78 |

**Table VI:**
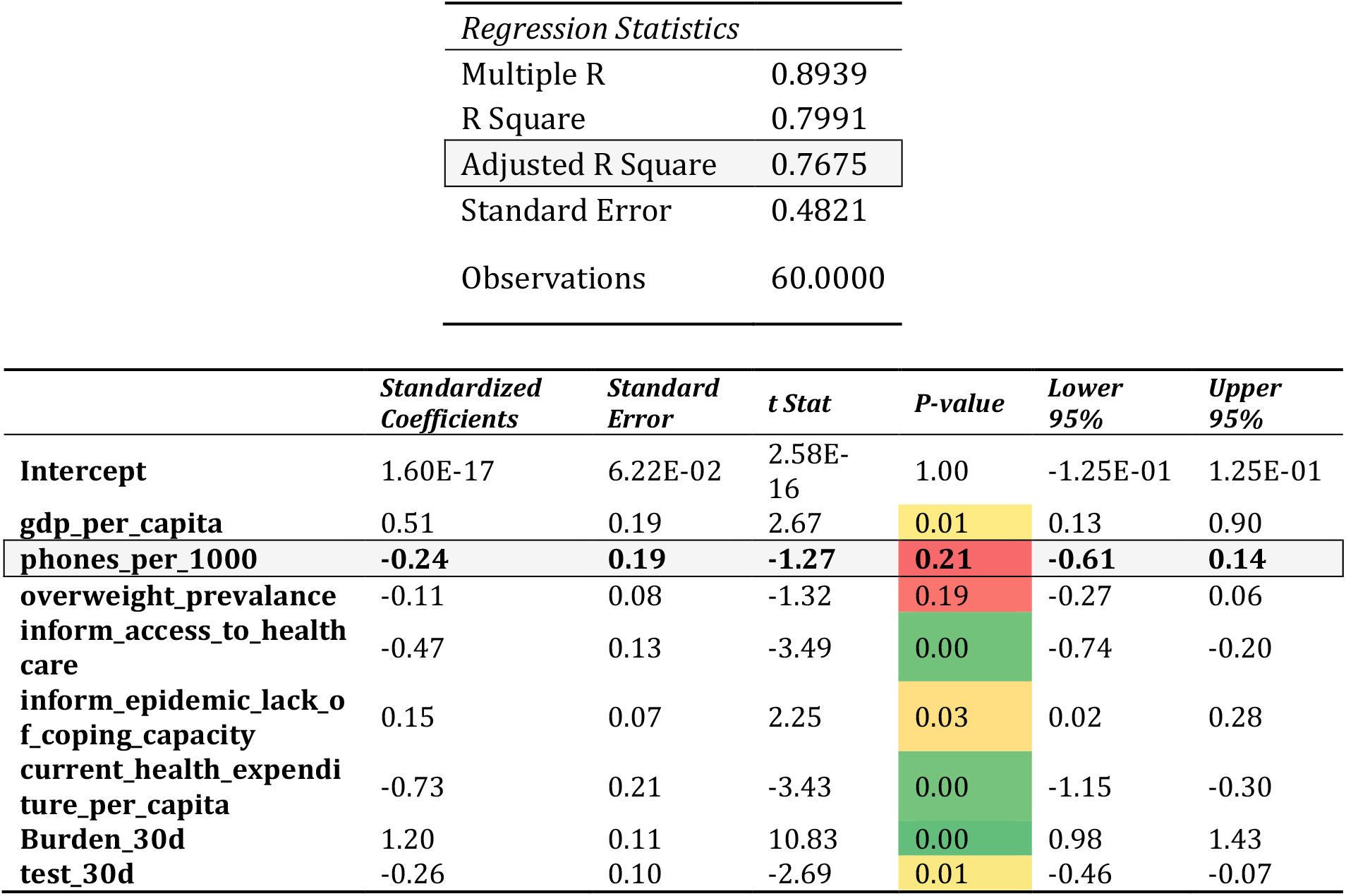
Best fitted model statistics for deaths.

| *Regression Statistics* |  |
| --- | --- |
| Multiple R | 0.8939 |
| R Square | 0.7991 |
| Adjusted R Square | 0.7675 |
| Standard Error | 0.4821 |
| Observations | 60.0000 |

|  | *Standardized Coefficients* | *Standard Error* | *t Stat* | *P-value* | *Lower 95%* | *Upper 95%* |
| --- | --- | --- | --- | --- | --- | --- |
| Intercept | 1.60E-17 | 6.22E-02 | 2.58E-16 | 1.00 | -1.25E-01 | 1.25E-01 |
| gdp_per_capita | 0.51 | 0.19 | 2.67 | 0.01 | 0.13 | 0.90 |
| phones_per_1000 | -0.24 | 0.19 | -1.27 | 0.21 | -0.61 | 0.14 |
| overweight_prevalance | -0.11 | 0.08 | -1.32 | 0.19 | -0.27 | 0.06 |
| inform_access_to_health care | -0.47 | 0.13 | -3.49 | 0.00 | -0.74 | -0.20 |
| inform_epidemic_lack_of_coping_capacity | 0.15 | 0.07 | 2.25 | 0.03 | 0.02 | 0.28 |
| current_health_expenditure_per_capita | -0.73 | 0.21 | -3.43 | 0.00 | -1.15 | -0.30 |
| Burden_30d | 1.20 | 0.11 | 10.83 | 0.00 | 0.98 | 1.43 |
| test_30d | -0.26 | 0.10 | -2.69 | 0.01 | -0.46 | -0.07 |

From **Table V**, we see that *burden_30d* can be best fitted by *phones_per_1000, agriculture, net_migration, overweight_prevalence, inform_access_to_healthcare, and current_health_ependiture_per_capita* while considering *test_30d* as a confounding factor. Among these factors, only the association between *net_migration, inform_access_to_healthcare, current_health_ependiture_per_capita* and *test_30d* are significant (p-values < 0.05). The overall indication is a bit counter-intuitive because the direction of association (the sign of the standardized coefficient) shows that the number of cases irrespective of tests performed increases with better healthcare access, higher health expenditure per capita and when more people are immigrating abroad. In reality, the burden of COVID-19 increased rapidly for countries with a better economic structure, such as many European countries and the USA. Most of those countries have excellent access to healthcare, and their health expenditure per capita is much higher than the rest of the world.

Also, it is understandable that the disease can spread to countries that have more tourists. However, many countries closed the borders at the early onset of the pandemic and therefore stopped the inward flow of foreign nationals. But that did not prevent the citizens of those countries living abroad to return home in large numbers due to the crisis. Hence, we found the direction of association of *net_migration* with cases to be reasonable. It indicates that considering all the other factors to be constants, including the number of tests performed, a country that has more returning citizens is likely to have more cases in comparison to countries with less number of citizens returning from abroad. The standardized beta coefficients indicate that after *test_30d*, the next most influential factor for the disease burden is the current per capita health expenditure (std. beta 0.55) followed by the access to healthcare rating (std. beta 0.37). The standard beta indicates that for one standard deviation increase/decrease of the independent variable, the dependent variable will increase/decrease by beta standard deviation if all the other factors remain constant. The sign of the coefficient indicates the direction of change.

The factors associated with the mortality rate is more intuitive. From **Table VI**, we see that most significantly associated factors with deaths are *gdp_per_capita, inform_access_to_healthcare, inform_epidemic_lack_of_coping_capacity, current_health_expenditure_per_capita, burden_30d* and *test_30d*. The model shows that countries with better access to healthcare, higher expenditure per capita for health, and better capacity to cope with epidemics can keep the number of deaths lower when all the other factors in the model are constant. The values of the standard beta coefficients indicate that after *burden_30d* the next most influential factor for *mortality_30d* is again the current per capita health expenditure (std. beta -0.73) followed by access to healthcare rating (std. beta -0.47). However, the direction of association is now inverse comparing to association with *burden_30d*. The fact that these two variables prevailed even after using *burden_30d* as a confounding factor indicates the strong association of these factors with *mortality_30d*.

A fascinating factor is *phones_per_1000*, which showed insignificant associations with the disease burden and mortality rates (p-value > 0.05). However, *phones_per_1000* was needed in both models for the best fit. From **Table V** and **Table VI**, we see that with increasing values of *phones_per_1000*, the number of cases increases. In contrast, the number of deaths decreases considering all the remaining factors are constant. On the one hand, it indicates that the disease spreads more rapidly in countries with more phone usage. It is possible because viruses are known to live longer and spread easily from touch-screen surfaces, and phones can easily be a medium to transfer germs from a surface to a person’s hand [38] [39]. On the other hand, a higher number of phones per 1000 is an indicator of a better communication system, easier access to healthcare information, and facilities for a country, all of which can play a major role in fighting COVID-19. Therefore, it is understandable that those countries handle the pandemic better by keeping the severity at bay. Smartphones are also considered to be used for contact tracing in many countries [40] [41]. Contact tracing is playing a pivotal role in educating the people, containing the disease, and taking early precautions for individuals with a high likelihood of being infected [42].

In **Figure 3** and **Figure 4**, we plotted the percentages of *burden_30d* among *test_30d* and the percentages of *mortality_30d* among the *burden_30d* for a few countries. The two measures provide us a relative idea about which countries are most affected and handle the pandemic better. The country names were sorted from high to low according to the percentages of burden among test values, and the top 15 and bottom 15 countries were chosen for these plots. For example, the burden to test ratio in the USA is moderately high (16.85%), according to **Figure 3**. Still, the mortality to burden ratio is low (2.67%), whereas Kenya has one of the lowest burden to test ratio (only 1.99%), but the mortality to burden ratio is quite high (4.38%). Ecuador is another good example with a very high burden to test ratio (50.78%) but moderate mortality to burden ratio (4.72%). It is worth mentioning that different countries have different criteria to perform a COVID-19 test on an individual. So, it is probable that the testing criteria might confound some of the independent variables. Converting testing criteria into numerical variables and including them as confounding factors can be a potential future research direction.

**Figure 3:**
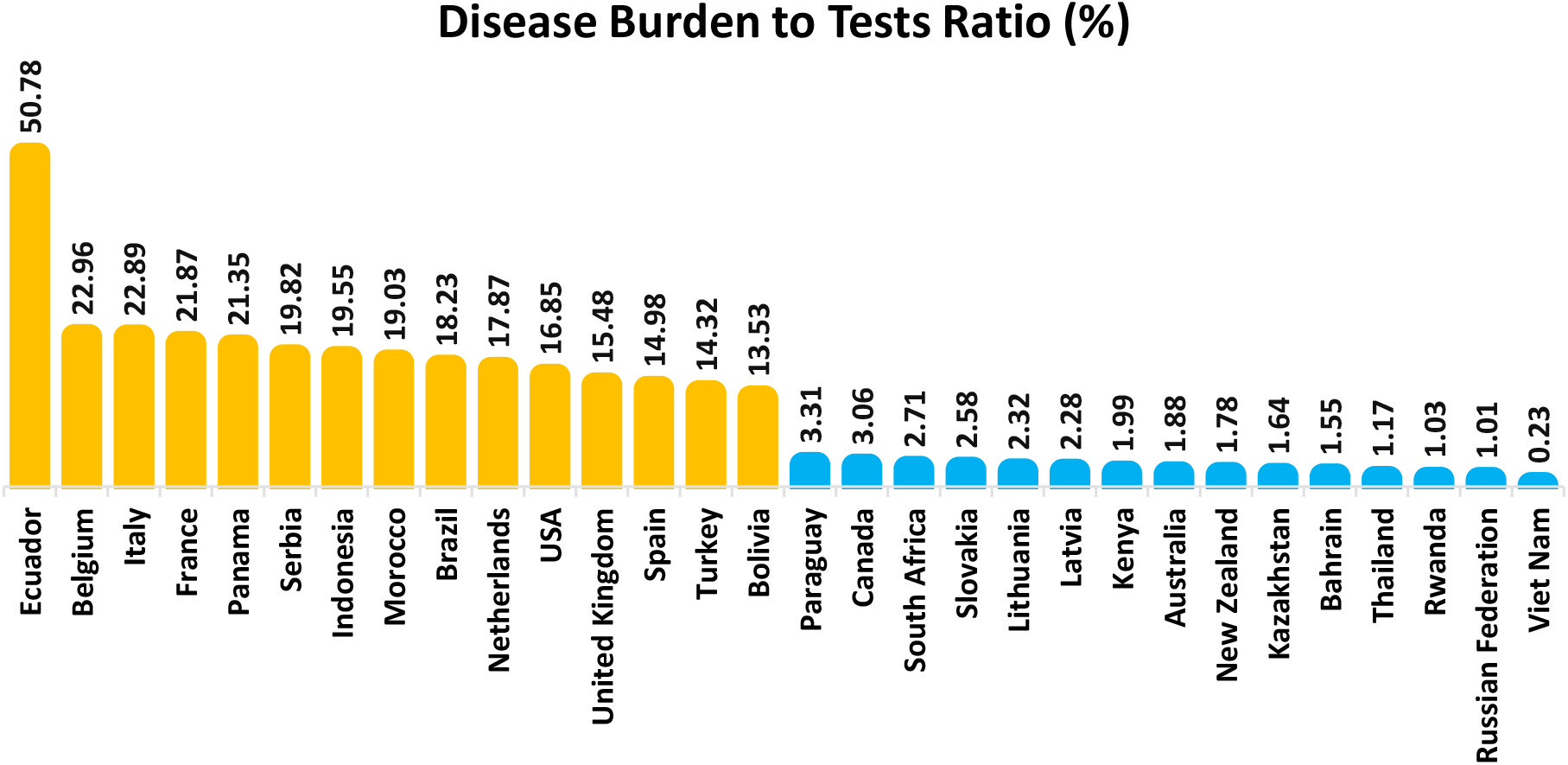
Bar plot showing the ratio of COVID-19 disease burden to the number of tests performed on the 30^th^ day from the first 20 reported cases for 30 selected countries. The percentage values are sorted, and countries with the top 15 (yellow) and the bottom 15 (blue) percentages are chosen for demonstration.

**Figure 4:**
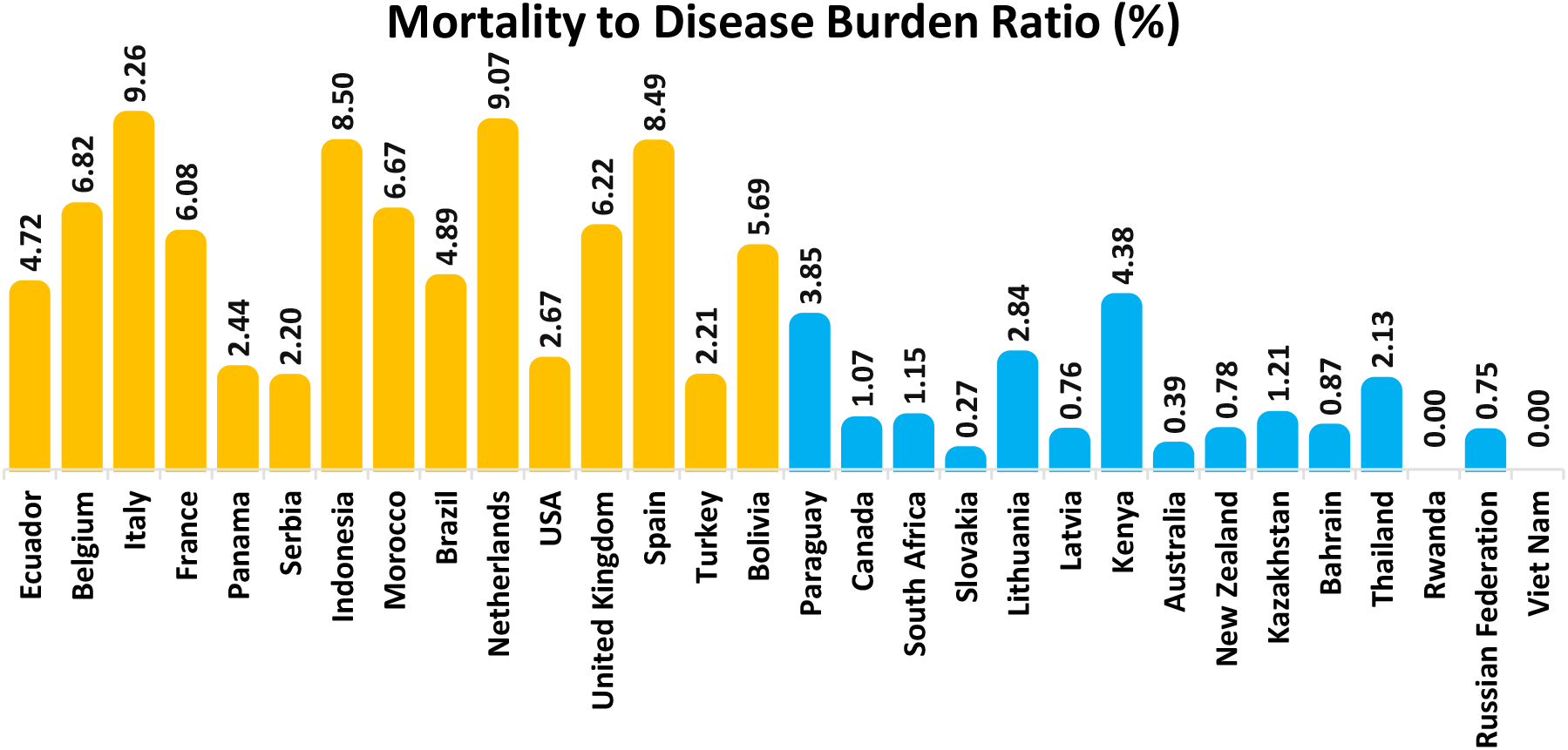
Bar plot showing the mortality to disease burden ratio (%) after the 30-day window for the same countries depicted in Figure 3.

We investigated the factors further and provided more insight in **Table VII** for a few countries with a very different socioeconomic and cultural scenario. In this table, we sorted the countries according to high to low severity, i.e., the mortality to burden ratio (%) measure, and analyzed the standardized factors associated with the *mortality rate* model. We color-coded the standardized values such that transition from red to green would indicate a decrease in *mortality_30d*. The table tells us that even though Italy has a modest health expenditure per capita compared to countries like Bangladesh or India, its lacking in coping capacity with an epidemic is playing a major role in its being severely affected by the pandemic. India and Bangladesh, on the other hand, have relatively lower healthcare expenditure per capita and therefore are facing moderately high mortality to disease burden ratio (%) even though their disease burden to test ratios (%) are smaller than both Italy and the USA. The USA is benefitting from the higher expenditure on health per capita and good pandemic coping capacity to keep the severity in check even though it has spread rapidly. There are some cases, such as Slovakia, for which it is not apparent why the country has a lower disease burden and mortality rate while it neither has a good coping capacity nor does it spend more on healthcare. There are likely some other factors dictating the disease burden and mortality in those countries – such as the government’s policy on the issue, cultural factors, efficiency of contact tracing, COVID-19 testing criteria, etc.

**Table VII:** **Standardized values of the best-fitting variables with standardized death for a few countries and the corresponding mortality to burden ratio (%) and burden to test ratio (%). Sorted by the mortality to burden ratios (%).**

| country | gdp_per_capita | phone_s_per_1000 | overweight_prevalance | inform_access_to_healthcare | inform_epidemic_lack_of_coping_capacity | current_health_expenditure_per_capita | Mortality to Disease Burden Ratio (%) | Disease Burden to Tests Ratio (%) |
| --- | --- | --- | --- | --- | --- | --- | --- | --- |
| Italy | 0.952 | 0.597 | 0.402 | -1.076 | 2.954 | 0.439 | 9.260 | 22.886 |
| Bangladesh | -1.098 | -1.439 | -2.441 | 1.374 | 0.395 | -1.133 | 3.705 | 10.259 |
| India | -1.015 | -1.256 | -2.454 | 1.579 | -0.319 | -1.062 | 2.805 | 3.707 |
| USA | 1.869 | 2.842 | 1.073 | -0.719 | -1.272 | 3.472 | 2.668 | 16.852 |
| Slovakia | -0.156 | -0.416 | 0.234 | -0.362 | 1.347 | -0.152 | 0.270 | 2.581 |

Finally, the results of our country-level aggregated analysis do not directly match with other studies like [23] because of several reasons. In [23], the authors considered the first 28 days after the first reported case of a country to find the disease burden and mortality rate. However, because of the testing criteria and government policies, many countries did not either perform the tests on a wide scale or report the cases in a structured and timely manner. Hence, there were even countries with only one reported case within that time frame. On the other hand, we took the 30 days after the first reported 20 total cases – which is a period when the pandemic started to spread rapidly, and we were able to capture the trend of the pandemic better. The cumulative cases ranged between 47 and 161679 (median 3257), and cumulative deaths went between 0 and 6803 (median 69). Also, our analysis includes health infrastructure-related factors, and some of those factors prevailed after the backward elimination indicating that from a country-level macro view, the burden and mortality rate of the disease is probably better described by such factors. On the other hand, most environmental, demographic, and population-level health factors got discarded

## Conclusion

In this paper, we provided a multivariate analysis to find relevant socioeconomic, environmental, and health-related factors influencing the COVID-19 pandemic’s disease burden and mortality rate across different countries of the world. We performed a correlation coefficient-based initial factor selection followed by a backward elimination-based systematic approach to find the best-fitted models with the disease burden and mortality count within a specific time window for a country with minimum human intervention. Our results show that even though the disease burden has increased more rapidly in economically affluent countries with a few exceptions, many such countries can keep the mortality rate in control. This phenomenon can be attributed to the better healthcare structure and higher expenditure for health per capita of those countries. Our results show a positive association of the disease burden with the emigration rate. We attributed it to the recent international border shutdown and return of citizens to their home countries in large numbers. We also found that countries with a higher density of phones have more massive disease burden but lesser mortality rate. The micro-level association of smartphone and digital devices with the spread of COVID-19 can thus be an interesting future research direction. On the other hand, a higher number of phones indicate affluence, and we have already found that more affluent countries are coping with the mortality rate better.

One limitation of this work is that we only considered country-level aggregated statistics, and therefore, the associations will be different if individual subjects are considered instead. It is understandable that for particular subjects, the factors would also be associated with individuals rather than an aggregated entity, such as a country. Backward elimination has some limitations, and, in many cases, it is probably preferable to hand-pick the variables. However, we resorted to backward elimination to provide a systematic approach for fitting instead of a subjective one. We acknowledge that it is probable to design a better-fitted model with a more extensive trial/error approach. However, considering that the outcomes that we obtained are reasonable and in-line with the general trend of the pandemic and intuitively understandable, we believe that our analysis is pointing us in the right direction. Poisson regression or Negative Binomial regression might be used to find better-fitted models instead of multiple linear regression. However, as explained in [37], the most statistically significant parameters of all these methods should theoretically be the same, and therefore we think our results and analysis would still hold. Finally, as we move deeper into the pandemic and include more countries and more reliable data, the relationships can be reevaluated to find even better models.

## Data Availability

All data used in this research are publicly available in the links below:
1. CSSE, JHU, "COVID-19 Data Repository by the Center for Systems Science and Engineering (CSSE) at Johns Hopkins University," 2020. [Online]. Available: https://github.com/CSSEGISandData/COVID-19. [Accessed 28 April 2020].
2. Our World in Data, "Data on COVID-19 (coronavirus) by Our World in Data," 2020. [Online]. Available: https://github.com/owid/covid-19-data/tree/master/public/data. [Accessed 28 April 2020].
3. ICAAP, "Data Sets," 15 Mar 2015. [Online]. Available: http://gsociology.icaap.org/dataupload.html. [Accessed 28 April 2020].
4. Berkeley Earth, "Climate Change: Earth Surface Temperature Data," 2017. [Online]. Available: https://www.kaggle.com/berkeleyearth/climate-change-earth-surface-temperature-data?select=GlobalLandTemperaturesByCountry.csv. [Accessed 28 April 2020].
5. The World Bank, "World Development Indicators," 2020. [Online]. Available: https://databank.worldbank.org/source/world-development-indicators. [Accessed 28 April 2020].
6. INFORM, "INFORM COVID-19 Risk Index Version 0.1.2," 2020. [Online]. Available: https://data.humdata.org/dataset/inform-covid-19-risk-index-version-0-1-2.
7. Roche Data Science Coalition, "UNCOVER COVID-19 Challenge," April 2020. [Online]. Available: https://www.kaggle.com/roche-data-science-coalition/uncover/version/3. [Accessed 28 April 2020].

